# Occurrence of relative bradycardia and relative tachycardia in individuals diagnosed with COVID-19

**DOI:** 10.1101/2022.02.02.22270342

**Authors:** Aravind Natarajan, Hao-Wei Su, Conor Heneghan

## Abstract

**Background:** The COVID-19 disease caused by the Severe Acute Respiratory Syndrome Coronavirus 2 (SARS-CoV-2) has become one of the worst global pandemics of the century causing tremendous human and economic suffering worldwide. While considered a respiratory disease, COVID-19 is known to cause cardiac complications. Wearable devices are well equipped to measure heart rate continuously and their popularity makes them valuable devices in the field of digital health. In this article, we use Fitbit devices to examine resting heart rate from individuals diagnosed with COVID-19 ^a^

**Methods:** The Fitbit COVID-19 survey was conducted from May 2020 - June 2021. We collected resting heart rate data from 7,200 individuals (6,606 symptomatic, 594 asymptomatic) diagnosed with COVID-19 between March 2020 - December 2020, as well as from 463 individuals diagnosed with influenza between January 2020 - December 2020. Data from healthy individuals served as a control, in order to model the seasonal variation. We also computed heart rate variability and respiratory rate data for symptomatic COVID-19.

**Findings:** Resting Heart Rate is elevated during COVID-19 symptom onset, with average peak increases relative to the baseline of 1.8%±0.1% (3.4%±0.2%) for females (males), where the quoted numbers are mean and standard error of the mean. After the initial peak, the resting heart rate decreased and reached a minimum on average *≈* 13 days after symptom onset. The minimum value relative to the baseline is more negative for females (−1.75% ± 0.1%) compared to males (0.08% ± 0.2%). The resting heart rate then increased, reaching a second peak on average *≈* 28 days from symptom onset, before falling back to the baseline *≈* 112 days from symptom onset. All estimates vary with disease severity.

**Interpretation:** The resting heart rate is modified for several months following a COVID-19 diagnosis. Interestingly, this effect is seen with seasonal influenza also, although the bradycardia minimum and the second tachycardia peak are often more pronounced in the case of symptomatic COVID-19. By computing resting heart rate daily, wearable devices can contribute to monitoring wellness during recovery from COVID-19, and seasonal influenza.

**Funding:** A.N., H.-W.S., and C.H. are supported by Fitbit Research, Google LLC.

**Research in Context:** *Evidence before this study:* We searched PubMed, Google, and Google Scholar for research articles published in English up to Oct 31, 2021, using common search terms such as “bradycardia and COVID-19”, “cardiac complications and COVID-19”, etc. Articles were also retrieved by searching through citations of known literature. It is known that COVID-19 can cause cardiac complications such as bradycardia and arrhythmias. Using data from commercially available wearable devices, it has been shown previously that the resting heart is elevated during symptom onset, then decreases reaching a minimum, before rising again to attain a second peak, before finally returning to the baseline.

*Added value of this study:* We present results from the largest (to our knowledge) dataset considered to-date, involving 7200 participants (6606 symptomatic and 594 asymptomatic) diagnosed with COVID-19. We also present results from 463 individuals diagnosed with influenza. Our large dataset allows us to perform more detailed examinations by age, disease severity, and sex. We also discuss the time evolution of heart rate variability and respiratory rate. The heart rate variability shows a similar time evolution as the resting heart rate but with opposite phase, while the respiratory rate decreases monotonously following the peak at symptom onset.

*Implications of all the available evidence:* The results presented in this work show that commercially available trackers and smart-watches can help in monitoring heart health in the weeks and months following a COVID-19 diagnosis. An estimate of the amplitude of the bradycardia dip may provide information valuable to critical care.

## I. INTRODUCTION

Cardiac complications are known to be associated with the COVID-19 disease caused by the novel Severe Acute Respiratory Syndrome - Coronavirus 2 (SARS-CoV-2)^1–8^. Cardiac injury, heart failure, and arrhythmias have been recorded in patients diagnosed with COVID-19^1,5,9,10^.

An unusual feature that is sometimes seen in patients diagnosed with COVID-19 is the appearance of bradycardia, i.e. slow heart rate, or heart rate not increasing as expected with body temperature^11–14^. Amaratunga et al.^15^ found bradycardia in a study of 4 patients with confirmed COVID-19, with minimum pulse rates in the range 42−49 beats per minute. Amir et al.^16^ reported 6 cases of bradycardia among patients diagnosed with COVID-19, with 4 patients developing complete atrioventricular block. Elikowski et al.^17^ presented clinical data of 19 patients diagnosed with COVID-19 who exhibited sinus bradycardia, in some cases showing heart rates as low as 32 bpm during daily hours. Srinivasan et al.^18^ discussed 6 cases of patients who were diagnosed with COVID-19 and admitted with normal sinus rhythm, and who subsequently developed sinus bradycardia with daytime heart rates ranging from 35−48 bpm. In a study of 97 patients with a non-severe presentation of COVID-19, Zhou et al.^19^ found significant sinus bradycardia (below 50 bpm) in 7.2% of cases. Guo et al.^20^ reported ventricular tachycardia (VT) or ventricular fibrillation (VF) in 11 out of 187 patients with confirmed COVID-19 and found that elevated levels of troponin T were correlated with VT/VF. Inflammatory damage due to cytokines has been suggested as a possible explanation for cardiac involvement with COVID-19^17,21–23^.

Commercially available wearable devices have been shown to be useful in early detection of COVID-19 and for monitoring symptoms^24–29^. Radin et al.^30^ studied resting heart rate (henceforth RHR) data from Fitbit devices to investigate long term changes following symptom onset. RHR is typically elevated around symptom onset. We use the term “relative” to indicate that the RHR is elevated/decreased relative to the baseline value for that individual, although the RHR is not necessarily above/below the clinical threshold guideline^31^. They also found the RHR exhibits a dip which we refer to as transient relative bradycardia provided the RHR is below the baseline value. The RHR dip was followed by a second elevated RHR peak. They found that the RHR was elevated for up to 79 days from symptom onset.

In this article, we obtain results consistent with the findings of Radin et al., and expand upon existing work in a number of ways. We study a much larger sample size than previously considered. We investigate how the resting heart range changes, for male and female individuals, and for individuals with severe, mild or asymptomatic presentations of COVID-19. We also consider individuals diagnosed with the seasonal influenza (henceforth “flu”). We tabulate the expected amplitudes of the maxima/minima, as well as the time taken to reach these maxima/minima, and the estimated widths of the peaks/troughs. We examine how these parameters vary with age, sex, and disease severity. We also study heart rate variability and respiratory rate and how these metrics vary with time.

## II. METHODS

### A. Survey Data

The Fitbit COVID-19 survey was conducted from May 21, 2020 to June 10, 2021, and collected data from participants residing in the USA or Canada. Participants provided information on whether they were diagnosed with COVID-19 or flu, as well as the test date, symptoms, and the start date of symptoms. Participants could optionally provide information about their age, sex, body mass index, and information on underlying conditions. Individuals diagnosed with COVID-19 also indicated the severity of the disease which could be (i) severe, indicating that they required hospitalization, (ii) mild, indicating that they recovered at home, or (iii) asymptomatic. The survey and associated marketing and recruitment materials were approved by an Institutional Review Board (Advarra). The participants provided electronic informed consent for their data to be used for research. In this study, we consider data from Fitbit users who reported testing positive for COVID-19 in the date range March 1 - Dec 31, 2020, as well as users who reported testing positive for flu in the date range Jan 1 - Dec 31, 2020. There were 11,918 participants who tested positive for COVID-19 (mean age = 40.8 yr, std. dev. = 12.4 yr, 79.0% female) in our dataset, and 865 participants who tested positive for flu (mean age = 41.7 yr, std. dev. = 13.3 yr, 78.2% female). We randomly select 1,000 users who did not report a positive test for either COVID-19 or flu as a control group (mean age = 45.3 yr, std. dev. = 13.9 yr, 71.6% female). Table I shows the prevalence of symptoms (self reported) for COVID-19 and flu, for male and female participants (for COVID-19, we also separate by disease severity). Some symptoms such as fatigue, headache and body ache are common for both COVID-19 and flu. By contrast, a decrease in taste and/or smell is more likely in the case of COVID-19 (72.6% female, 59.8% male) compared to flu (21.2% female, 11.6% male). Fever is more common with flu (81.3% female, 76.8% male) compared to COVID-19 (51.4% female, 58% male). Fig. 1 shows the distribution of positive cases for flu and COVID-19, where the horizontal axis is the test date, and only positive cases are shown. Cases of flu peaked in March 2020, while COVID-19 cases peaked much later.

**TABLE I.** Prevalence of symptoms, for COVID-19 (mild/severe) and flu, for male and female participants.

| Symptom | Mild (fem.) | Mild (male) | Severe (fem.) | Severe (male) | Flu (fem.) | Flu (male) |
| --- | --- | --- | --- | --- | --- | --- |
| Fatigue | 0.847 | 0.761 | 0.885 | 0.816 | 0.843 | 0.689 |
| Headache | 0.793 | 0.640 | 0.789 | 0.618 | 0.661 | 0.415 |
| Decrease in taste/smell | 0.727 | 0.598 | 0.701 | 0.592 | 0.211 | 0.116 |
| Congestion | 0.715 | 0.571 | 0.503 | 0.342 | 0.541 | 0.512 |
| Body ache | 0.689 | 0.666 | 0.793 | 0.684 | 0.818 | 0.677 |
| Cough | 0.652 | 0.627 | 0.784 | 0.730 | 0.731 | 0.616 |
| Chills | 0.532 | 0.557 | 0.713 | 0.632 | 0.734 | 0.604 |
| Fever | 0.501 | 0.563 | 0.749 | 0.796 | 0.813 | 0.768 |
| Sore throat | 0.498 | 0.385 | 0.418 | 0.342 | 0.560 | 0.378 |
| Loss of appetite | 0.472 | 0.345 | 0.701 | 0.638 | 0.440 | 0.293 |
| Shortness of breath | 0.432 | 0.347 | 0.871 | 0.836 | 0.396 | 0.299 |
| Chest pain | 0.401 | 0.284 | 0.687 | 0.533 | 0.334 | 0.244 |
| Diarrhea | 0.399 | 0.319 | 0.524 | 0.454 | 0.192 | 0.189 |
| Neck pain | 0.296 | 0.186 | 0.322 | 0.191 | 0.193 | 0.116 |
| Hoarse voice | 0.280 | 0.179 | 0.320 | 0.303 | 0.279 | 0.110 |
| Eye pain | 0.232 | 0.161 | 0.232 | 0.197 | 0.106 | 0.067 |
| Stomach ache | 0.225 | 0.115 | 0.290 | 0.118 | 0.130 | 0.104 |
| Confusion | 0.178 | 0.135 | 0.368 | 0.349 | 0.114 | 0.140 |
| Vomiting | 0.089 | 0.044 | 0.271 | 0.118 | 0.115 | 0.067 |
| Rash | 0.067 | 0.034 | 0.131 | 0.066 | 0.029 | 0.037 |
| Swelling in fingers/toes | 0.044 | 0.022 | 0.133 | 0.079 | 0.032 | 0.043 |

**FIG. 1.**
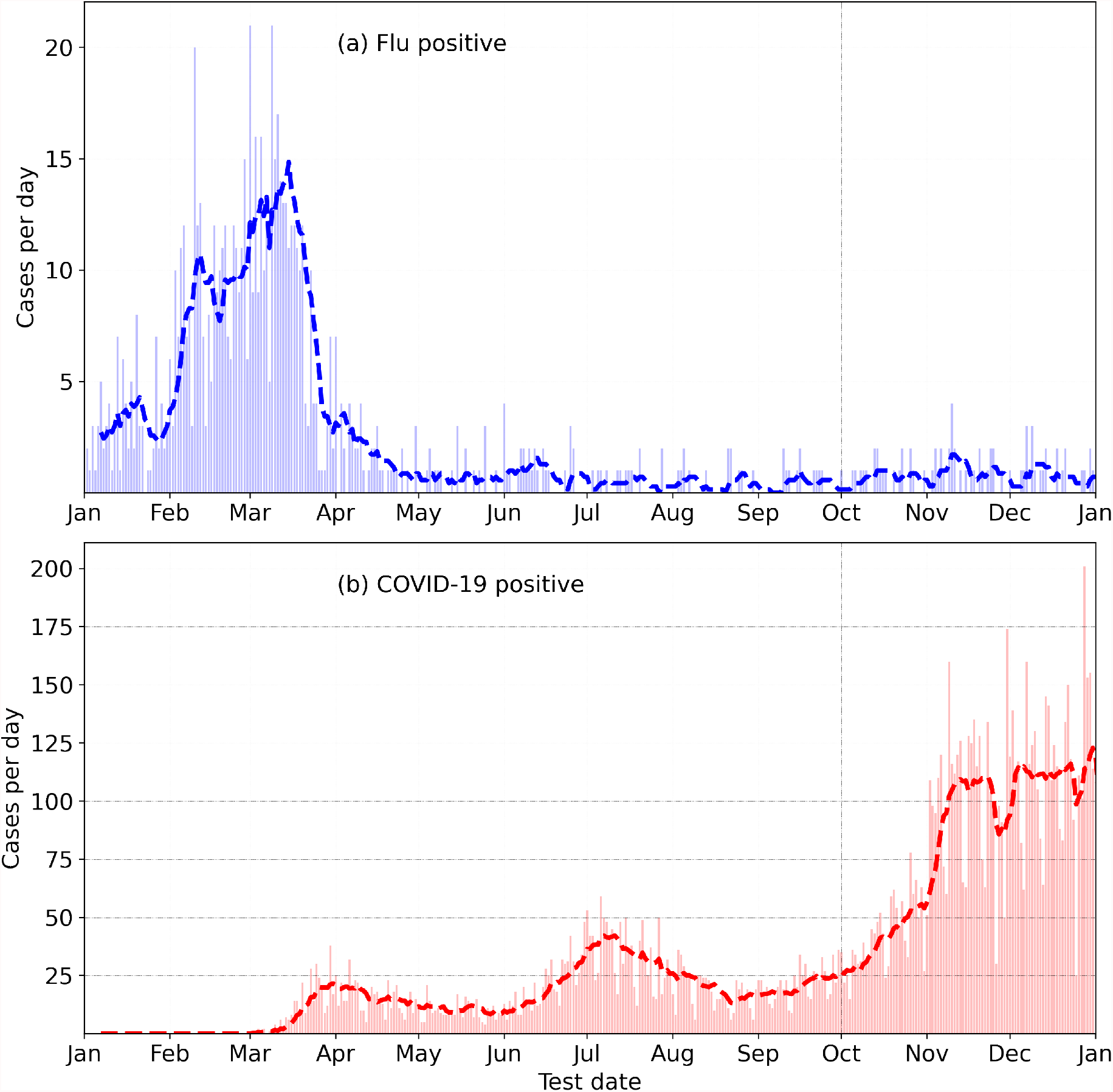
Incidence of Flu (top) and COVID-19 (bottom) in the year 2020, from the Fitbit COVID-19 survey.

### B. RHR data

We measure the RHR from participants in our study, collected using Fitbit devices. The “resting heart rate” as defined by Fitbit is the value closest to the heart rate measured when lying down just before waking up in the morning^32^.

RHR is computed using heart rate data during sleep when sleep data is available, and a proprietary algorithm is utilized to predict the RHR from the time series heart rate data during sleep. If sleep data is unavailable, RHR is computed using wake time heart rate data, at times when no activity is detected. The RHR is also processed with a Kalman filter which serves to smooth the waveform. Note that the RHR is not the minimum value of heart rate.

### C. Time variation of the RHR data

Let *D*_0_ be the date of symptom onset for symptomatic individuals (and the test date for asymptomatic individuals). Thus for symptomatic individuals, *D*_−1_ is one day prior to the appearance of symptoms, and *D*_+1_ is one day post appearance of symptoms. We computed the mean value ⟨*RHR*⟩ for each individual by averaging the RHR values from *D*_−90_ to *D*_−15_. We discard data from participants who have fewer than 30 measurements in this time window. With this data sufficiency condition, our dataset contains 463 individuals with the flu, and 7200 individuals with COVID-19 (6606 symptomatic, 594 asymptomatic). We compute the fractional change in RHR *ξ* from days *D*_−14_ to *D*_+180_ as:

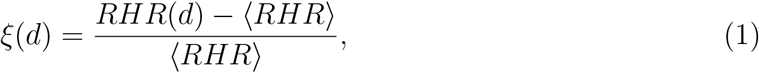

where *d* is a variable indicating day index.

### D. Controlling for the seasonality of resting heart rate

The RHR has a known seasonal modulation. In a study of 200,000 individuals wearing Fitbit devices and residing in the United States, Quer et al.^33^ found a change in the population’s average RHR by 2 beats per minute (bpm). In the Northern hemisphere, the RHR peaks in the first week of January and reaches its minimum at the end of July.

Since all participants in our study reside in the USA or Canada, we assume that they are subject to the same seasonal trends. We use the control group to estimate how RHR varies with the time of year, by applying Eq. 1 to obtain *ξ*_control_ as follows:

1. Randomly sample a date from the COVID-19 or flu distribution and set to *D*_0_ (date of symptom onset).
2. Compute *ξ*_control_(*d*) using Eq. 1 from the control group for dates *d* relative to *D*_0_. We may then subtract out the seasonality to find the effect of illness on RHR:

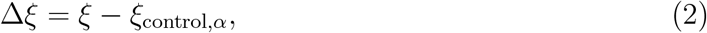

where *α* may be flu or COVID-19. Note that *ξ*_control_ must be computed twice: once for the COVID-19 distribution, and once for the flu distribution. To compute Δ*ξ*(*d*) for a given individual, we would use the appropriate *ξ*_control,*α*_ depending on whether the participant was diagnosed with flu or COVID-19.

### E. Estimation of parameters

We estimate a number of parameters from the time variation of Δ*ξ*. These are summarized in Table II. The waveforms for *ξ* and *ξ*_control_ are linearly interpolated with a step size of 0.2 days. The waveforms are then smoothed with a Savitzky−Golay filter with a window length of 7 days. We compute Δ*ξ* from the smoothed waveforms, and estimate the peaks/troughs by means of a peak detection algorithm. We also report an estimated error on the parameter values using the jackknife technique^34,35^.

**TABLE II.**
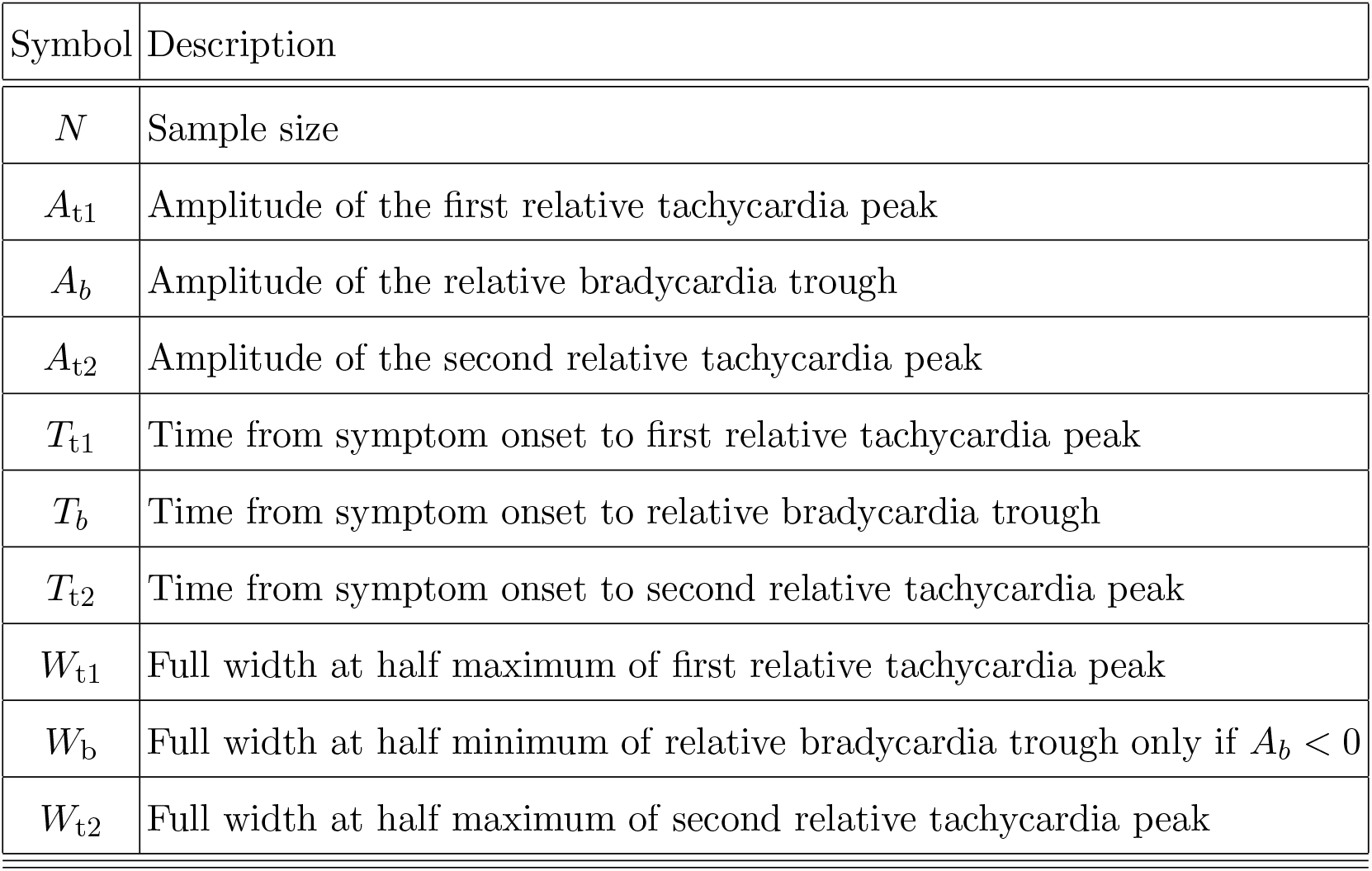
Description of parameters.

| Symbol | Description |
| --- | --- |
| $N$ | Sample size |
| $A_{t1}$ | Amplitude of the first relative tachycardia peak |
| $A_b$ | Amplitude of the relative bradycardia trough |
| $A_{t2}$ | Amplitude of the second relative tachycardia peak |
| $T_{t1}$ | Time from symptom onset to first relative tachycardia peak |
| $T_b$ | Time from symptom onset to relative bradycardia trough |
| $T_{t2}$ | Time from symptom onset to second relative tachycardia peak |
| $W_{t1}$ | Full width at half maximum of first relative tachycardia peak |
| $W_b$ | Full width at half minimum of relative bradycardia trough only if $A_b < 0$ |
| $W_{t2}$ | Full width at half maximum of second relative tachycardia peak |

### F. Heart Rate Variability and Respiratory Rate

Details on how Fitbit devices compute respiratory rate and heart rate variability have been previously published^28,36^. In this article, we consider RMSSD computed in 5 minute windows between the hours of midnight and 6 am. The median of the measurements is then reported. For the respiratory rate calculation, we first compute the power spectral density of interbeat intervals in 5 minute windows between the hours of midnight and 7 am. The measurements are then averaged, and the respiratory rate is estimated from the averaged power spectral density. We subtract out the seasonality of the RMSSD and respiratory rate in a similar manner to the RHR, with the addition of a median filter (with a period of 7 days). The median filter was necessary because the RMSSD and respiratory rate waveforms are not smoothed by a Kalman filter, and are hence noisier than the RHR.

### G. Code

All computations are performed using standard Python libraries.

## III. RESULTS

### A. Change in RHR following a Flu / COVID-19 diagnosis

Fig. 2 shows the fractional change in RHR for flu and COVID-19, for symptomatic individuals. Subplot (a) compares *ξ* and *ξ*_control_ for flu, while subplot (b) compares *ξ* and *ξ*_control_ for COVID-19. The horizontal axis *D*_*n*_ is the date relative to the date when symptoms first appear (*n* < 0 are prior to symptom onset, while *n >* 0 are after symptom onset. *D*_0_ is the date when symptoms first present). *ξ* and *ξ*_control_ are averaged over all individuals for a specific *D*_*n*_. Subplot (c) shows Δ*ξ*, i.e. the excess fractional change in RHR after the seasonal variation has been subtracted. From subplot (c), we infer the following:

**FIG. 2.**
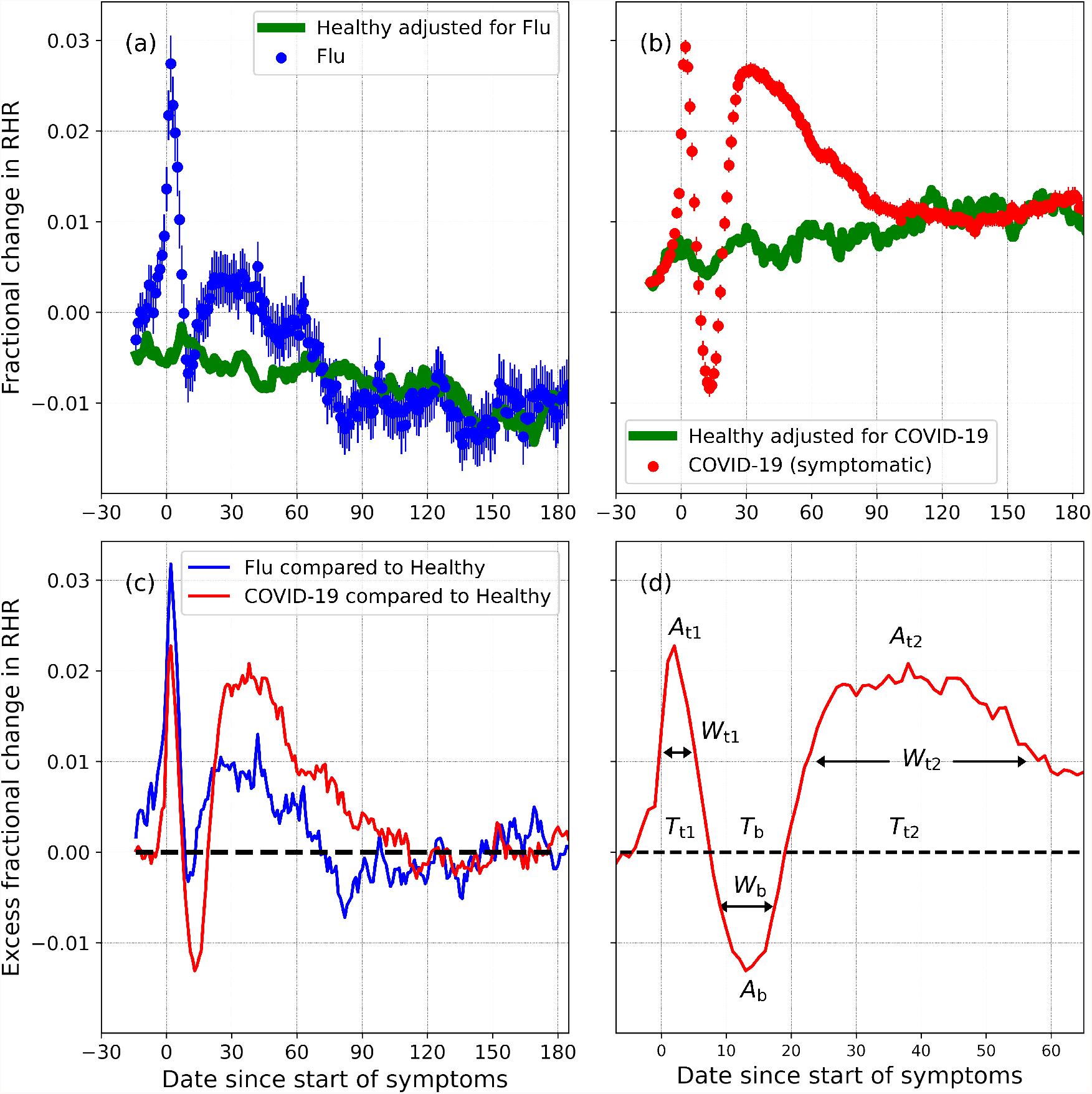
Fractional change in RHR (*ξ*), for individuals diagnosed with flu (a) and COVID-19 (b), along with the expected seasonal variation (*ξ*_control_). (c) shows the excess fractional change in RHR (Δ*ξ*) for flu and COVID-19. The parameters shown in (d) are defined in Table II.

1. The excess fractional change in RHR, i.e. Δ*ξ* is elevated on average during the onset of symptoms. Δ*ξ* reaches a peak value *A*_t1_ at a time *T*_t1_ days from the onset of symptoms. This is the first transient relative tachycardia.
2. Following the peak, Δ*ξ* decreases, reaching a minimum value *A*_b_ at a time *T*_b_ days from the onset of symptoms. If *A*_b_ < 0, we refer to the trough as transient relative bradycardia.
3. Following the minimum, Δ*ξ* increases again, reaching a second peak *A*_t2_ at a time *T*_t2_ days from symptom onset. This is the second transient relative tachycardia.
4. Past *T*_t2_, Δ*ξ* decreases and eventually falls to zero, indicating that the RHR variation is no longer due to illness.

The parameters *A*_t1_, *A*_b_, *A*_t2_, *T*_t1_, *T*_b_, *T*_t2_, *W*_t1_, *W*_b_, and *W*_t2_ are marked in subplot (d), and described in Table II.

Fig. 3 shows the excess fractional change in RHR (Δ*ξ*) for COVID-19 and flu, for varying severity, and for male and female individuals. Subplot (a) shows the effect of severity on Δ*ξ*. The magenta curve is plotted for severe cases (i.e cases that required hospitalization). The brown curve shows mild cases, while the green curve is plotted for asymptomatic cases. The blue curve shows Δ*ξ* for flu. The amplitudes of the two relative tachycardia peals *A*_t1_ and *A*_t2_ are much larger in the case of severe COVID-19. The bradycardia dip (*A*_b_) is more pronounced in the case of mild COVID-19. The first relative tachycardia peak amplitude (*A*_t1_) is larger for flu compared to mild or asymptomatic COVID-19, while the second tachycardia peak is similar in the cases of flu and asymptomatic COVID-19. For the cases of severe, mild, asymptomatic, and flu respectively, we find Δ*ξ* falls to zero approximately 118, 112, 79, and 71 days after the onset of symptoms (or test date for asymptomatic cases).

**FIG. 3.**
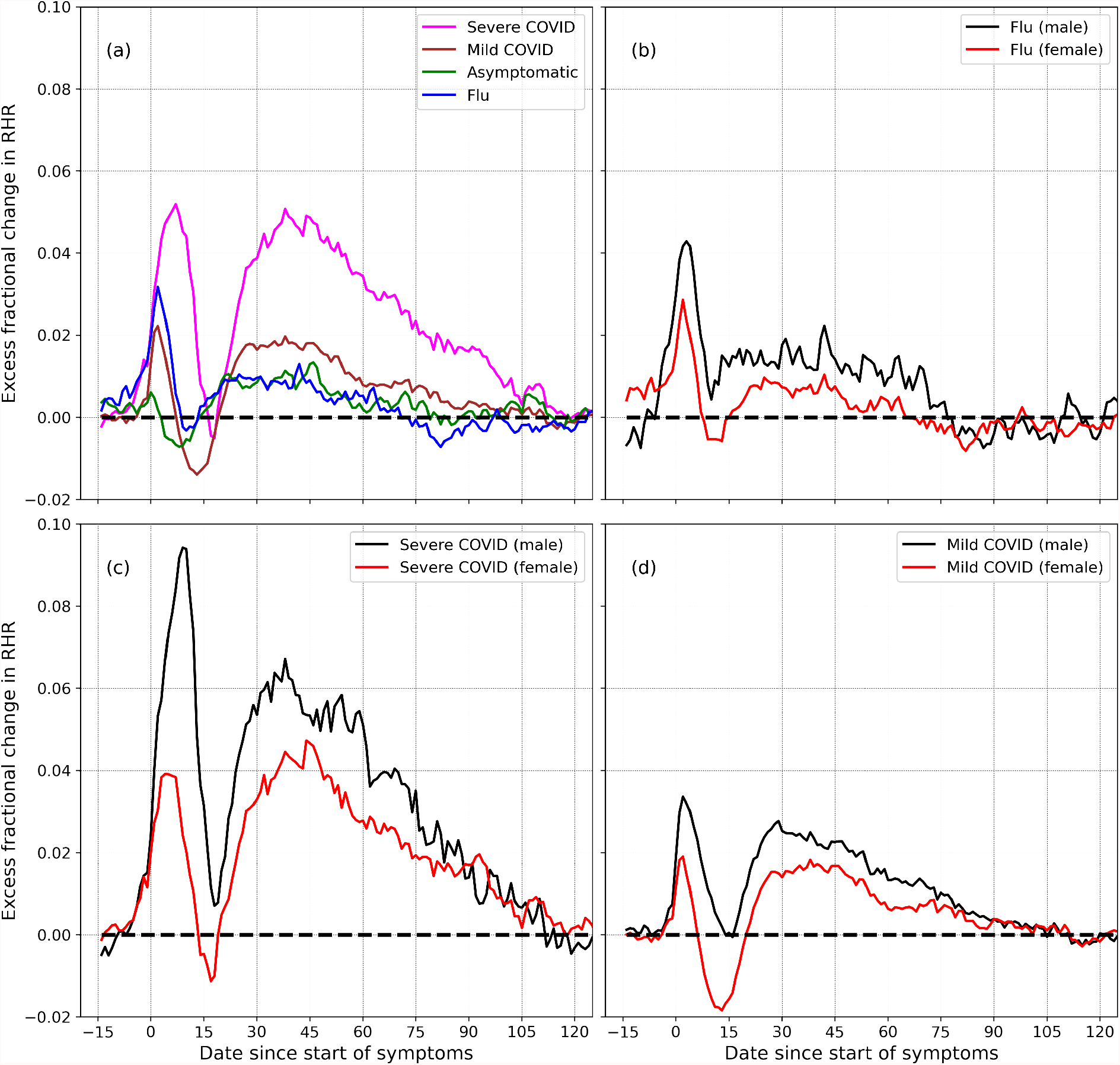
Excess fractional change in RHR (Δ*ξ*), variation with severity and sex: (a) shows Δ*ξ* for severe, mild, and asymptomatic COVID-19, as well as flu. (b) shows Δ*ξ* for male and female individuals diagnosed with flu. (c) and (d) show Δ*ξ* for male and female participants, for the cases for severe and mild COVID-19 respectively.

Subplot (b) shows the difference between male and female individuals who were diagnosed with flu. Similar to the case of COVID-19, the peak amplitudes *A*_t1_ and *A*_t2_ are higher for males compared to females. The trough amplitude *A*_*b*_ is lower for females. Subplots (c) and (d) shows the variation of Δ*ξ* for male and female individuals, for severe and mild COVID-19 cases respectively. In both cases, the peak amplitudes *A*_t1_ and *A*_t2_ are higher for males, for both severe and mild cases of COVID-19. The minimum amplitude *A*_b_ on the other hand is lower in females than in males, and is more negative for mild cases of COVID-19 compared to severe cases.

Table III shows the estimated values of the amplitudes for the RHR minimum (*A*_b_) and the two maxima (*A*_t1_ and *A*_t2_), along with jackknife estimated standard errors. Table IV shows the estimated values of the various parameters, for female and male participants, and for different ages. Each peak/trough is characterized by 3 parameters: the amplitude, time from symptom onset, and full width at half maximum/minimum. The 2 relative tachycardia peaks are parameterized by (*A*_t1_, *T*_*t*1_, *W*_t1_) and (*A*_t2_, *T*_*t*2_, *W*_t2_), while the minimum is parameterized by (*A*_b_, *T*_b_, *W*_b_). Note that *W*_b_ is only defined when *A*_b_ < 0 indicating relative bradycardia. Table V shows the estimated values of the parameters for male and female participants, for severe, mild, and asymptomatic COVID-19 presentations, as well as for flu. Fig. 4 shows the estimated mean values of these parameters for different age groups, for male (black) and female (red) individuals.

**TABLE III.** Estimation of peak amplitudes. *N* is the sample size. Estimates are mean and jackknife standard error of the mean.

| Disease | Sex | $N$ | $A_{t1}$ | $A_b$ | $A_{t2}$ |
| --- | --- | --- | --- | --- | --- |
| COVID-19 | F | 5183 | $1.78 \pm 0.08$ | $-1.75 \pm 0.10$ | $1.81 \pm 0.08$ |
| COVID-19 | M | 1411 | $3.39 \pm 0.18$ | $0.08 \pm 0.18$ | $2.80 \pm 0.17$ |
| Flu | F | 360 | $2.60 \pm 0.32$ | $-0.57 \pm 0.33$ | $0.96 \pm 0.30$ |
| Flu | M | 103 | $4.28 \pm 0.67$ | $0.66 \pm 0.72$ | $1.94 \pm 0.58$ |

**FIG. 4.**
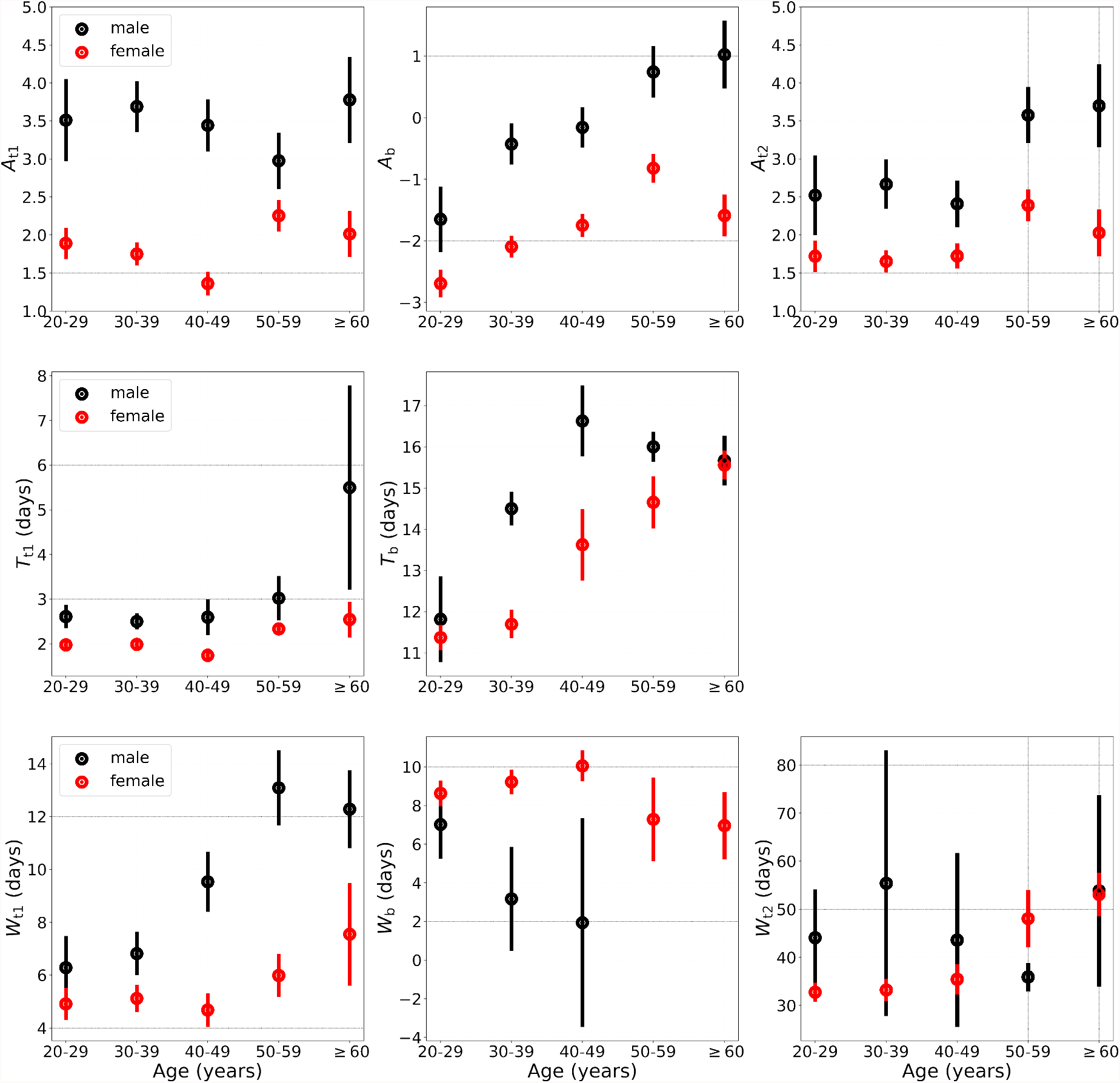
Parameters describing the time evolution of the RHR, from Table V, for different age groups, for male (black) and female (red) individuals. Jackknife error bars represent the standard error of the mean. Subplot for *T*_t2_ is not shown.

### B. Relative bradycardia and relative tachycardia

Let us now quantify the prevalence of relative bradycardia and relative tachycardia. To do this, we define the following 4 windows each of which are 15 days long (starting and ending dates inclusive, and *D*_0_ is the date when symptoms present, only symptomatic individuals included):

- “Control” window from *D*_−45_ to *D*_−31_.
- Presumed “healthy” window from *D*_−30_ to *D*_−16_.
- Presumed “relative bradycardia” window from *D*_+7_ to *D*_+21_.
- Presumed “relative tachycardia” window from *D*_+22_ to *D*_+36_.

The healthy and control windows are expected to contain data when participants are healthy. The RHR is averaged over each window, and we only consider participants who have 15 days of data in each window. Let ΔRHR_*w*_ be the difference between the RHR averaged over window *w* and the RHR averaged over the control window:

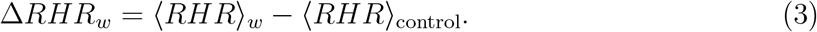

We compute the fraction of participants who have ΔRHR *≤ x* bpm, computed from the bradycardia window, and compare it to the same fraction computed from data in the healthy window, where *x* is a threshold value plotted along the horizontal axis. We do a similar comparison with the data in the tachycardia window. We compute the fraction of participants with ΔRHR *≥ x* computed from the tachycardia window, and compare with the fraction computed from data in the healthy window. Fig. 5 shows this comparison. The curve plotted in red in Fig. 5(a) shows the fraction of individuals in the time window *D*_+7_ − *D*_+21_ with ΔRHR *≤ x* plotted along the horizontal axis. This should be compared with the curve plotted in green which is an estimate of how likely it is that such a low ΔRHR may occur due to random chance. Fig. 5(b) shows a similar comparison, but for ΔRHR *≥ x*, and considering the tachycardia window.

**FIG. 5.**
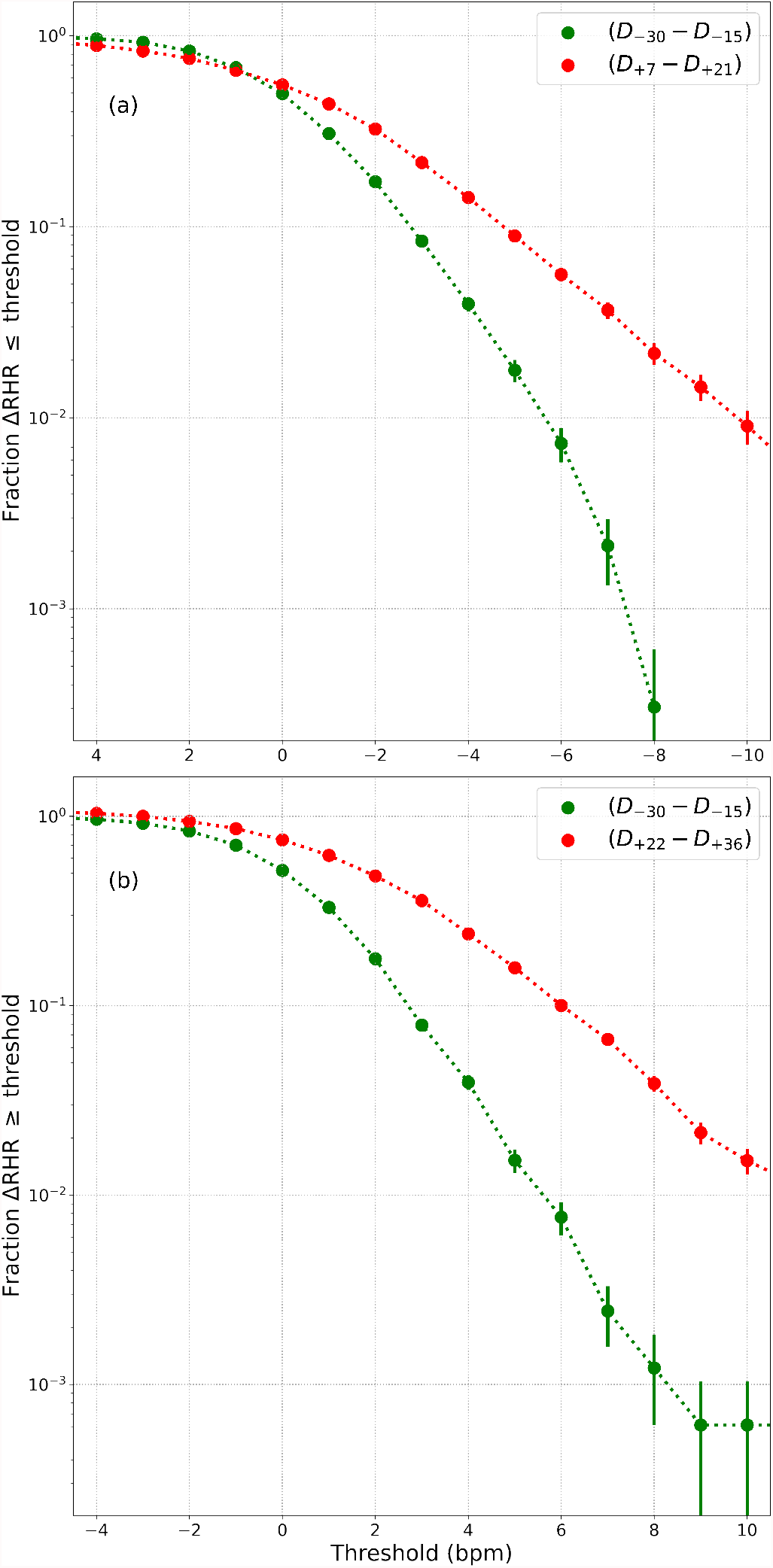
(a) Fraction of individuals with ΔRHR *≤ x* in the relative bradycardia window (red points), where *x* is a threshold, compared to the fraction that could be expected due to chance (green points). (b) shows the fraction of individuals with ΔRHR *≥ x* in the relative tachycardia window (red points), compared to the fraction that could be expected due to chance (green points). ΔRHR is the excess RHR compared to the control window. Error bars show standard error of the mean.

Interestingly, there is a correlation between the peak value of ΔRHR measured during symptom onset (we use a 15 day window from *D*_−5_ to *D*_+9_) and the peak and minimum values of ΔRHR measured in the second relative tachycardia window (*D*_+22_ to *D*_+36_) and the relative bradycardia window (*D*_+7_ to *D*_+21_). Fig. 6 shows this correlation. The horizontal axis is ΔRHR during symptom onset, while the vertical axis shows ΔRHR during either the second tachycardia phase (red) or the bradycardia phase (blue). We see that ΔRHR_tachy_ measured in the second relative tachycardia phase is positively correlated with ΔRHR_symptom_, the peak value observed at symptom onset. Similarly, ΔRHR_brady_, the minimum value observed during the relative bradycardia phase is positively correlated with ΔRHR_symptom_. This means that a low ΔRHR at symptom onset is indicative of a decreased RHR in the relative bradycardia phase. An approximate estimate of ΔRHR_tachy_ and ΔRHR_brady_ may be obtained by the following linear relations (with all measurements in bpm):

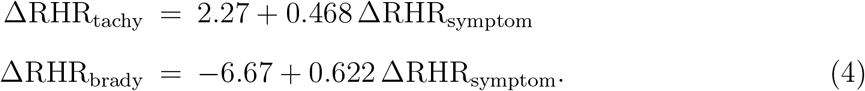

**FIG. 6.**
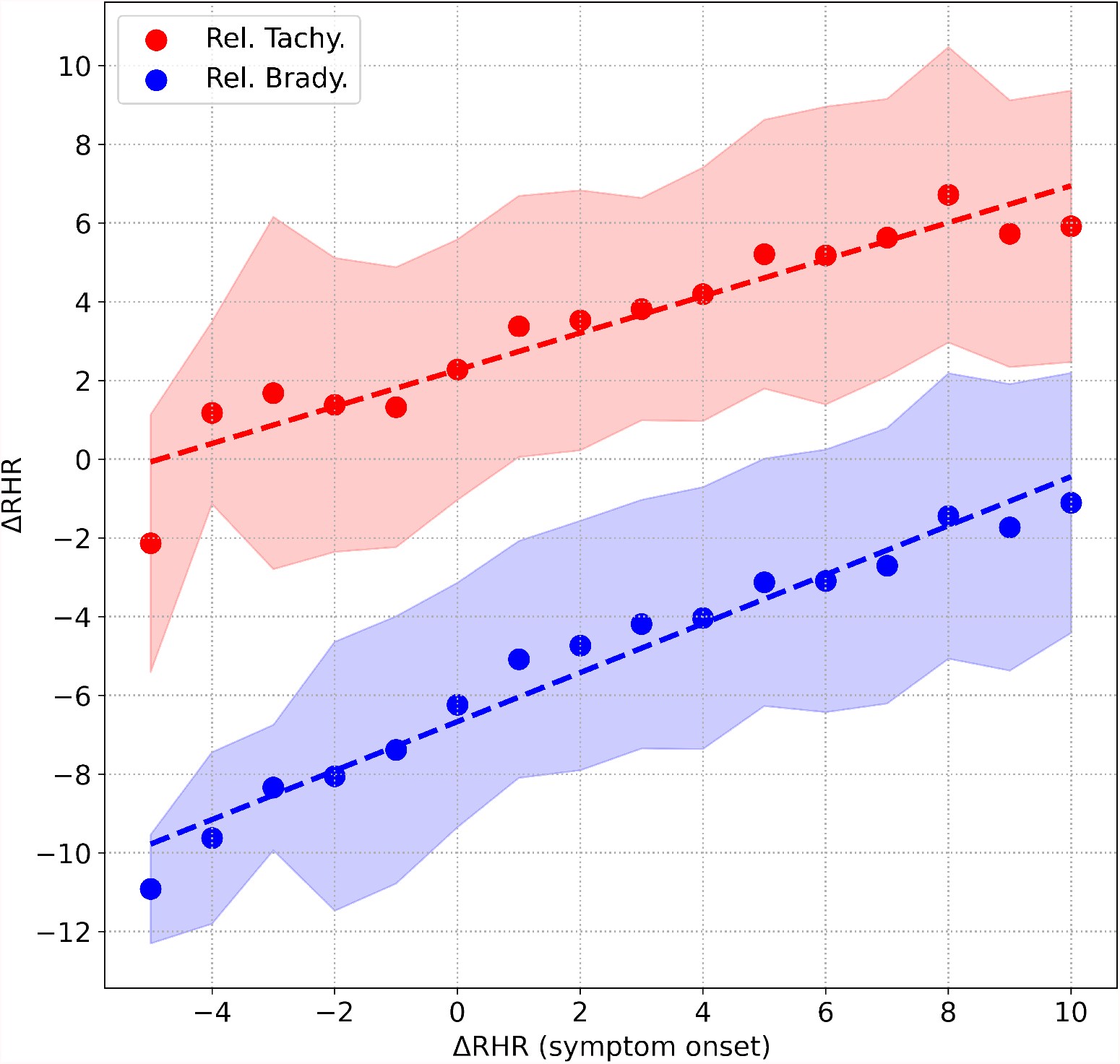
Correlation between the peak value of ΔRHR measured during symptom onset, and (i) peak value of ΔRHR in the second relative tachycardia window shown in red, (ii) minimum value of ΔRHR measured in the relative bradycardia window shown in blue. The shaded areas represent the 1 standard deviation range.

### C. Comparison with other measurements

Let us now briefly consider 2 more health metrics: the heart rate variability quantified by the root mean square of the successive differences (RMSSD) and the respiratory rate. Fig. 7 shows the excess fractional change Δ*ξ* in RMSSD (subplot (a)) and the respiratory rate (subplot (b)), plotted for symptomatic COVID-19. Also shown for comparison in dashed lines is the fractional change in RHR. The fractional change in RMSSD shows a similar time evolution as the RHR, except with opposite phase, i.e. when the RHR is elevated, the RMSSD is decreased and vice-versa. The relative bradycardia phase is seen to be accompanied by elevated heart rate variability, while the second relative tachycardia phase is associated with reduced heart rate variability. The variation is also of greater amplitude with the RMSSD compared to the RHR. The fractional change in the respiratory rate, on the other hand, does not show a similar time evolution. The respiratory rate is elevated during symptom onset, in response to the illness. It then decreases sharply, and then gradually returns to normal. Thus there is no equivalent of the relative bradycardia and second relative tachycardia phases with the respiratory rate.

**FIG. 7.**
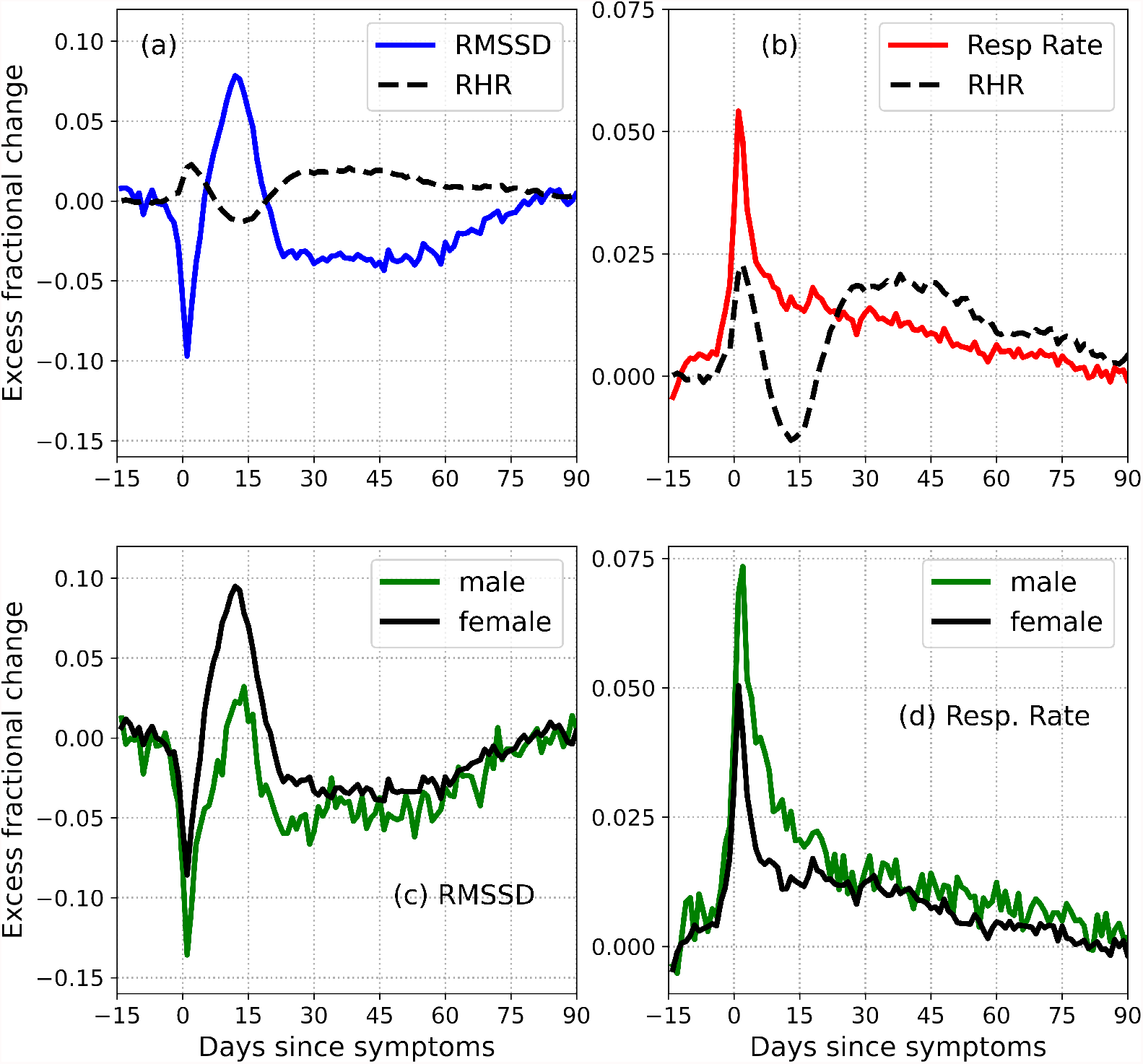
Excess fractional change in RMSSD and respiratory rate. Subplots (a) and (b) also show the excess fractional change in RHR (dashed black lines). Note that the RHR waveform is smoothed over time, resulting in a more rounded peak. Subplots (c) and (d) show the RMSSD and respiratory rate for male (green) and female (black) subjects respectively.

## IV. DISCUSSION

Both COVID-19 and flu cases show 3 distinct phases:

- First relative tachycardia phase during symptom onset when the RHR is elevated above normal, reaching a local peak value *A*_t1_ at a time *T*_t1_ days after symptom onset, with a full width at half maximum *W*_t1_. The peak value *A*_t1_ is higher in males compared to females, for COVID-19 (*p*−value < 0.0001) and flu (*p*−value = 0.0118).
- Following this peak, the RHR decreases, and reaches a local minimum value *A*_b_ at a time *T*_b_ days after symptom onset. If *A*_b_ < 0, we refer to this phase as relative bradycardia, and define a full width at half minimum *W*_b_. The condition *A*_b_ < 0 is satisfied for females (averaged over all ages), for COVID-19 (*p* < 0.0001), and for flu (*p* = 0.042). The condition *A*_b_ < 0 is not statistically significant for males (averaged over all ages), for either COVID-19 or flu.
- Following the minimum, the RHR increases, reaching a second local maximum *A*_t2_ at a time *T*_t2_ days from symptom onset, with a full width at half maximum *W*_t2_. The peak value *A*_t2_ is higher in males compared to females, for COVID-19 (*p* < 0.0001) and flu (*p* = 0.0667).

Fig. 3 shows Δ*ξ* for male and female participants for different cases of COVID-19 severity, and for flu. Since these phases occur with both COVID-19 and flu, they are not novel to COVID-19. They can however, be more prominent in the case of COVID-19. It should be noted that flu has also been associated with cardiovascular disease^37^. It is also known that the autonomic nervous system plays a key role in sensing and responding to infection, with pathogens activating vagal signaling causing bradyarrhythmias^38^. It is intriguing that we see a statistically significant difference between male and female individuals, in the time variation of the RHR (Fig. 3) and the time variation of the RMSSD (Fig. 7).

The parameters that describe the time evolution of the RHR depend on sex, age, and disease severity and are tabulated in Tables III, IV and V. They are also plotted in Fig. 4 for different age groups for male and female individuals. The relative bradycardia minimum *A*_b_ and the time to the minimum *T*_b_ are correlated with age (The Pearson *r* for the correlation between *A*_b_ and age was found to be *r* = 0.80 for females and *r* = 0.97 for males. The Pearson *r* for the correlation between *T*_b_ and age was found to be *r* = 0.98 for females and *r* = 0.77 for males). The width of the first relative tachycardia peak *W*_t1_ is also correlated with age (*r* = 0.83 for females, and *r* = 0.94 for males).

**TABLE IV.** Estimation of parameters, by age. *Values of *T*_t2_ are only approximate due to noise and the large width of the second relative tachycardia peak.

| Age (yr) | Sex | $N$ | $A_{t1}$ | $A_b$ | $A_{t2}$ | $T_{t1}$ | $T_b$ | $T_{t2}^*$ | $W_{t1}$ | $W_b$ | $W_{t2}$ |
| --- | --- | --- | --- | --- | --- | --- | --- | --- | --- | --- | --- |
| 20-29 | F | 994 | 1.9 | -2.7 | 1.6 | 2 | 11.4 | 25.8 | 5 | 8.8 | 32.8 |
| 30-39 | F | 1627 | 1.8 | -2.1 | 1.7 | 2 | 11.6 | 27.4 | 5 | 9.4 | 33.2 |
| 40-49 | F | 1305 | 1.4 | -1.8 | 1.6 | 1.8 | 12 | 27 | 4.8 | 10 | 35.6 |
| 50-59 | F | 842 | 2.3 | -0.8 | 2.4 | 2.4 | 14.6 | 37.8 | 6 | 7.4 | 48 |
| $\geq 60$ | F | 364 | 2 | -1.6 | 2 | 2.6 | 15.6 | 38.6 | 7.6 | 6.8 | 53 |
| 20-29 | M | 178 | 3.5 | -1.6 | 2.5 | 2.6 | 11.8 | 33.8 | 6.4 | 7 | 44 |
| 30-39 | M | 392 | 3.7 | -0.4 | 2.7 | 2.4 | 14.2 | 27.6 | 6.8 | 3.2 | 55.6 |
| 40-49 | M | 370 | 3.4 | -0.2 | 2.4 | 2.6 | 16.6 | 28.8 | 9.4 | 2 | 43.8 |
| 50-59 | M | 275 | 3 | 0.6 | 3.7 | 3 | 17 | 28.8 | 13.2 | - | 36 |
| $\geq 60$ | M | 175 | 3.8 | 1 | 3.5 | 5.6 | 15.6 | 27.6 | 12.4 | - | 53.8 |

**TABLE V.**
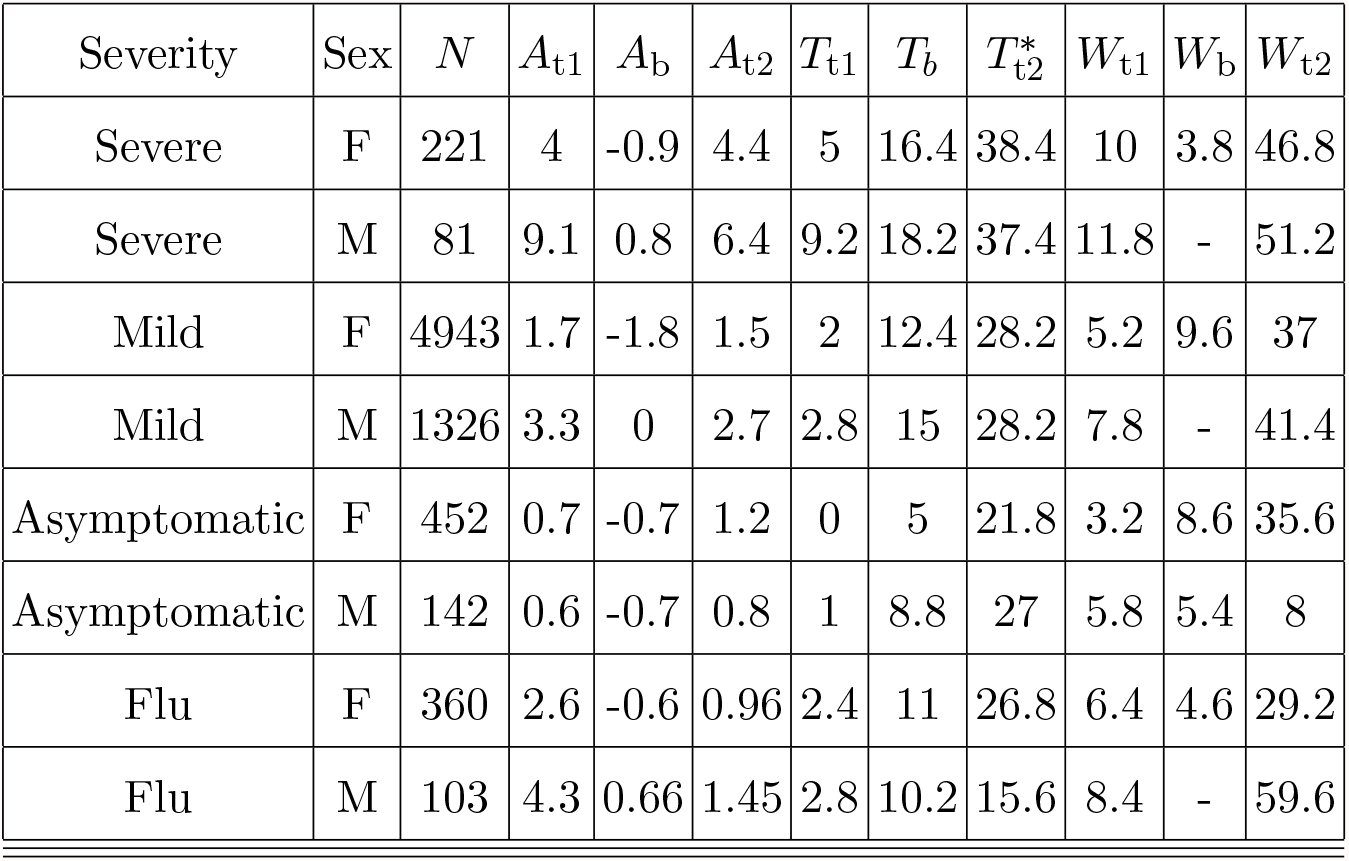
Estimation of parameters, by severity. *Values of *T*_t2_ are only approximate due to noise and the large width of the second relative tachycardia peak.

| Severity | Sex | $N$ | $A_{t1}$ | $A_b$ | $A_{t2}$ | $T_{t1}$ | $T_b$ | $T_{t2}^*$ | $W_{t1}$ | $W_b$ | $W_{t2}$ |
| --- | --- | --- | --- | --- | --- | --- | --- | --- | --- | --- | --- |
| Severe | F | 221 | 4 | -0.9 | 4.4 | 5 | 16.4 | 38.4 | 10 | 3.8 | 46.8 |
| Severe | M | 81 | 9.1 | 0.8 | 6.4 | 9.2 | 18.2 | 37.4 | 11.8 | - | 51.2 |
| Mild | F | 4943 | 1.7 | -1.8 | 1.5 | 2 | 12.4 | 28.2 | 5.2 | 9.6 | 37 |
| Mild | M | 1326 | 3.3 | 0 | 2.7 | 2.8 | 15 | 28.2 | 7.8 | - | 41.4 |
| Asymptomatic | F | 452 | 0.7 | -0.7 | 1.2 | 0 | 5 | 21.8 | 3.2 | 8.6 | 35.6 |
| Asymptomatic | M | 142 | 0.6 | -0.7 | 0.8 | 1 | 8.8 | 27 | 5.8 | 5.4 | 8 |
| Flu | F | 360 | 2.6 | -0.6 | 0.96 | 2.4 | 11 | 26.8 | 6.4 | 4.6 | 29.2 |
| Flu | M | 103 | 4.3 | 0.66 | 1.45 | 2.8 | 10.2 | 15.6 | 8.4 | - | 59.6 |

Among the 3 phases, the relative tachycardia at symptom onset is the easiest to understand, and may be interpreted as an immune response to infection. More unexpected is the decrease in RHR, possibly resulting in transient relative bradycardia. Approximately 1% of individuals experience a value of RHR that is at least 10 bpm lower than normal (see Fig. 5). Sinus bradycardia is known to occur during sleep in the case of COVID-19^39^. Since Fitbit’s measurement of RHR is not the lowest value of heart rate, this raises the possibility that the heart rate during sleep may reach dangerously low values. It is crucial to be aware of this transient relative bradycardia phase, since certain medications used in the treatment of COVID-19 such as Remdesivir have been known to cause bradycardia^40–42^. Interestingly, mild cases of COVID-19 are more likely to present with relative bradycardia. It is also more likely to present in females compared to males. A possible explanation for this may be obtained from Fig. 6, and Table I. Fig. 6 shows that the peak RHR during symptom onset is positively correlated with the magnitude of the dip, i.e. a smaller peak RHR during symptom onset is associated with a larger (and possibly negative) dip. It is well known the heart rate is strongly linked to temperature, increasing by 8.5 bpm per 1 degree C increase in temperature, as found in one study^43^. As seen from Table I, males are more likely to present with a fever compared to females. Similarly, fever is more common in severe cases compared to mild cases. Thus, females as well as mild cases presenting with a small RHR increase (or no increase) are more likely to experience a bradycardia dip compared to males, or severe cases. Similarly, Fig. 6 shows that the peak RHR during symptom onset is positively correlated with the height of the second tachycardia peak. As a result, the second tachycardia peak is higher in males, and for severe cases. The heart rate variability shows a similar time evolution as the resting heart rate, except with opposite phase. Fig. 7 shows the time evolution of the RMSSD (subplot (a)) and the respiratory rate (subplot (b)) with time. Unlike the RMSSD, the respiratory rate decreases monotonously past the initial peak. Subplots (c) and (d) show the variation of RMSSD and respiratory rate, for male and female subjects.

There are several limitations to the current work. A major concern is that we do not know whether the individuals who were diagnosed with COVID-19 and flu received medications to treat their illness, and what effects those medications may have had on their heart rate. The symptoms, severity, as well as symptom start date were all obtained from a survey, and could not be independently verified. We have also assumed that the participants were healthy prior to being diagnosed with COVID-19 or flu, and that the control group consisted of healthy individuals. Despite these limitations, the work presented in this article shows how COVID-19 and flu can alter the resting heart rate in the weeks/months following infection. This knowledge can be crucial in the case of patients with cardiac complications, or when patients are being treated with medication known to have cardiac effects.

## Data Availability

Fitbit's privacy policy does not permit us to make the raw data or aggregate data available to third parties including researchers.

## V. CONTRIBUTORS

A.N. and H.W.-S. had access to all the data and verified the results shown. A.N. was responsible for part of the scientific analysis and for preparing the first version of the manuscript. H.W.-S. was responsible for part of the scientific analysis. C.H. led the Fitbit COVID-19 study, and provided valuable insights and feedback on the analysis and presentation of results.

## VII. DATA SHARING

Our consent form and the protocol allow us to make the raw data available to two academic members of our research consortium, namely Stanford University, and Scripps Translational Research Institute. Fitbit’s privacy policy does not permit us to make the raw data or aggregate data available to third parties including researchers, outside of our web API Oauth 2.0 consent process. For specific questions, contact Fitbit at https://healthsolutions.fitbit.com/contact/.

## VIII. ACKNOWLEDGMENTS

A.N., H.W.-S. and C.H. acknowledge funding from Fitbit Research, Google LLC. We thank members of the Fitbit Research team for many helpful discussions.

## Footnotes

a This feature is not intended to diagnose or treat any medical condition and should not be relied on for any medical purposes. It is provided to Fitbit users to help manage well-being.

